# Implementation and early outcomes of a dedicated diabetes and hypertension clinic in rural Haiti: A 24-month retrospective cohort study

**DOI:** 10.64898/2026.07.22.26358732

**Authors:** Keddy Moise, Philippe Larco, Ernst David Charlotin, Kevin Mexis, Eddy Jean-Baptiste, Nancy Charles Larco

## Abstract

Noncommunicable diseases (NCDs), particularly diabetes and hypertension, are a growing cause of death in low-income countries. In Haiti, cardiovascular disease has overtaken HIV as the leading cause of adult death, and the isolated Grand’Anse Department had no dedicated chronic disease facility before this initiative. This retrospective single-center cohort study analyzed 1,298 encounters among 590 patients at a dedicated diabetes and hypertension clinic (the Centre Dr. René Charles) in Jérémie, Grand’Anse, from April 2024 to March 2026. Standardized assessments included serial blood pressure, fasting glucose, HbA1c, lipids, renal indices, and electrocardiography. Longitudinal change was modeled with linear mixed-effects models on calendar time, supplemented by paired first-to-last comparisons. The cohort was 74.1% female (mean age, 58.2 years), and nearly two thirds had primary-level education or none. Diagnoses were hypertension alone (52.9%), diabetes with hypertension (26.8%), and diabetes alone (20.3%). Establishing the clinic in this setting proved feasible, but 54% of patients attended only once, making early attrition the central finding. Among the minority who returned, exploratory within-patient analyses showed improvement in blood pressure: in mixed-effects models on calendar time, systolic pressure fell by 0.78 mmHg per month among patients with hypertension (95% CI, 0.36 to 1.21; P=0.005), the paired first-to-last decline was 9.0 mmHg (P<0.001), and control rose from 21.4% to 38.1% (P<0.001). These estimates come only from returners and are best read as hypothesis-generating. Glycemic control did not durably improve: despite a large paired fasting-glucose fall (52.5 mg/dL) and a borderline time trend (P=0.05), mean HbA1c remained 10.4%, with only 13.2% at target. Indication- driven screening of a clinically selected minority frequently detected end-organ damage, reflecting selective testing rather than cohort-wide prevalence. A dedicated NCD clinic can be established in one of the world’s most resource-limited settings; the priorities now are patient retention, broader medication access, and durable health-system investment.

## INTRODUCTION

Noncommunicable diseases (NCDs) cause an estimated 41 million deaths each year, and more than three quarters of these deaths occur in low- and middle-income countries (LMICs), where health systems built for episodic, acute care are poorly equipped for the sustained management that chronic disease demands.^1–4^ In sub-Saharan Africa, the share of deaths attributable to NCDs has risen over the past two decades, outpacing the capacity of health systems to respond.^5,6^ The Caribbean carries a particularly heavy burden: NCDs account for an estimated three quarters of all deaths in the region, driven by high rates of hypertension and a diabetes prevalence that has risen consistently over two decades.^7–9^ Across LMICs, patients are often lost to follow-up at every step of the care cascade. In a study of 44 countries, fewer than 40% of adults with hypertension had been diagnosed and only about 10% had achieved blood-pressure control; comparable gaps exist for diabetes.^10–13^

Haiti confronts this epidemic with one of the least-resourced health systems in the world, a gross domestic product of roughly 1,420 US dollars per capita (in constant 2015 dollars), and most of its population living below the national poverty line.^14,15^ Cardiovascular disease has overtaken HIV as the leading cause of adult death, yet it has drawn little of the programmatic investment directed for decades at communicable disease.^16,17^ The population-based Haiti Cardiovascular Disease Cohort found that roughly 30% of adults in Port-au-Prince had hypertension, of whom fewer than half were treated and about 13% were controlled,^18^ while the PREDIAH survey previously reported hypertension in nearly half of adults and in more than two thirds of those older than 40.^19,20^ Studies in Léogâne and Port-au-Prince report hypertension prevalence between 20% and 44%,^21,22^ and population-based data show an age-standardized chronic kidney disease prevalence of 14%, with diabetes and hypertension as the strongest correlates.^23^ The health system operates with approximately 2.5 physicians and 3.0 nurses per 10,000 population, and successive earthquakes and political crises have repeatedly destroyed facilities and disrupted supply chains, including across the southern peninsula.^24,25^ For most Haitians outside the capital, the regular follow-up, continuous medication supply, and laboratory monitoring that chronic disease care requires have been effectively out of reach.

The Grand’Anse Department, on the Tiburon peninsula of southwestern Haiti, is among the most isolated and underserved regions of the country. Its roughly 468,000 residents are dispersed across mountainous terrain with limited road access, and the departmental capital, Jérémie, has the only general hospital.^26^ Before the initiative reported here, the department had no chronic disease registry, screening protocol, or follow-up system, and patients with diabetes or hypertension were seen only episodically during acute illness. Experience elsewhere indicates that this gap can be closed. The World Health Organization Package of Essential NCD Interventions and the Chronic Care Model define feasible, protocol-driven approaches for primary care,^27–29^ and the HIV programs that reached treatment targets in these same fragile settings have shown that longitudinal chronic care is achievable when purpose-built infrastructure, standardized protocols, and trained staff are in place.^30,31^

In April 2024, the Fondation Haïtienne de Diabète et de Maladies Cardiovasculaires (FHADIMAC), with the support of the Sanofi Global Health Unit, opened the Centre Dr. René Charles in Jérémie, a dedicated ambulatory clinic for the systematic screening, diagnosis, and longitudinal management of diabetes and hypertension and the first facility of its kind in the Grand’Anse Department. Here we report the clinic’s first 24 months of operation. We describe enrollment, patient demographics, and clinical profiles; evaluate within-patient changes in blood pressure and glycemia across serial visits; quantify the burden of end-organ complications detected through targeted screening; and assess patient retention and its relation to clinical outcomes.

## METHODS

### Study Design and Setting

This retrospective, single-center cohort study evaluated the implementation of the Centre Dr. René Charles, an ambulatory chronic disease clinic in Jérémie, the departmental capital of Grand’Anse, Haiti, using routinely collected data from its clinical registry. The clinic receives patients from primary health centers, community health-worker referrals, and community-based screening campaigns. The study period spans the clinic’s inception in April 2024 through March 2026 (24 months). The study population comprised all adults presenting with type 1 or type 2 diabetes mellitus, hypertension, or both. Patients were classified by primary diagnosis into three mutually exclusive categories on the basis of their dossier coding: hypertension only (HT), diabetes mellitus with concurrent hypertension (DT), or diabetes mellitus without hypertension (DM).

The National Bioethics Committee of Haiti (Comité National de Bioéthique, Ministère de la Santé Publique et de la Population) reviewed and approved the study protocol and authorized the analysis (reference 2526-32), granting a waiver of written individual consent, given the low-literacy context of the population and the minimal-risk, non-interventional, registry-based design; verbal consent for inclusion in the registry was obtained from each patient at enrollment by the treating clinician, in Haitian Creole, and recorded in the clinical record. The registry was accessed for this retrospective analysis on 14 July 2026. As the clinic’s treating clinicians, the authors had access to identifying information in the registry during routine care; the analysis dataset was de-identified by removing direct identifiers (patient names) before analysis. The work was conducted in accordance with the principles of the Declaration of Helsinki. Reporting follows the Strengthening the Reporting of Observational Studies in Epidemiology (STROBE) guideline for cohort studies.^32^

### Clinical Assessments

At each encounter, trained clinical staff performed a standardized assessment. Blood pressure was measured with a calibrated manual sphygmomanometer and appropriate cuff size; a single seated reading was recorded after at least five minutes of rest. Anthropometric measures included weight, height, body-mass index (BMI), and waist circumference. Fasting blood glucose was measured by capillary point-of-care glucometer; when a patient had already eaten, a postprandial value was recorded instead and was not included in the fasting-glucose analysis. Laboratory investigations, performed on a scheduled basis and as clinically indicated, included glycated hemoglobin (HbA1c), a fasting lipid panel, serum creatinine with estimated glomerular filtration rate (eGFR, by the 2021 CKD-EPI equation without race adjustment), and spot urine albumin-to-creatinine ratio (ACR). Screening for end-organ damage included direct fundoscopy and 12-lead electrocardiography. Medications, behavioral risk factors, and sociodemographic data were recorded.

### Definitions

Blood-pressure control was defined as systolic pressure below 140 mmHg and diastolic pressure below 90 mmHg. Glycemic targets were HbA1c below 7.0% and fasting glucose below 126 mg/dL. BMI categories followed World Health Organization definitions, and abdominal obesity was defined as a waist circumference of at least 102 cm in men and 88 cm in women. Albuminuria was defined as an ACR of at least 30 mg/g, and chronic kidney disease of stage 3 or higher as an eGFR below 60 mL/min/1.73 m².

### Statistical Analysis

Analyses were performed in R, version 4.4.2 (R Foundation for Statistical Computing), using the readxl, dplyr, lme4, lmerTest, ggplot2, and patchwork packages. Continuous variables are summarized as means with standard deviations (SD) and categorical variables as counts with percentages. The primary longitudinal analysis modeled change in each cardiometabolic outcome over calendar time using linear mixed-effects models, with time measured as days since the patient’s first visit (expressed per month) as a fixed effect and a random intercept and random slope for time per patient; the model was reduced to a random intercept where the slope variance was not estimable. This approach uses all available measurements, accommodates the variable intervals between visits, and accounts for within-patient correlation and informative gaps in testing better than analyses indexed to ordinal visit number. Blood-pressure models were fit among patients with hypertension (HT or DT) and glycemic models among patients with diabetes (DM or DT). Fixed-effect slopes are reported per month with 95% Wald confidence intervals; degrees of freedom and P values use the Satterthwaite approximation.

As a clinically intuitive supplement, paired within-patient changes from the first to the most recent measurement were also computed. Mean paired changes are reported with 95% confidence intervals from paired t-tests, the change in blood-pressure control status with McNemar’s test, and proportions with Wilson-score 95% confidence intervals. Patient retention was characterized by the proportion returning for two or more visits, by time-horizon retention (the proportion of patients enrolled at least N days before study close who returned within N days, for N of 90, 180, and 365 days), and by median follow-up duration. Diagnostic coverage was reported as the proportion of eligible patients tested at least once for each investigation, because testing was not uniform. All analyses used available-case denominators with no imputation, and all tests were two-sided at an alpha level of 0.05. The four cardiometabolic outcomes were the pre-specified primary analyses, interpreted as hypothesis-generating given the single-arm observational design; all other analyses are secondary and unadjusted for multiplicity. The analyses followed a pre-specified statistical analysis plan (S1 File), and the full analysis and figure code is provided as Supporting Information (S2 File and S3 File).

## RESULTS

### Enrollment and Utilization

Between April 2024 and March 2026, the clinic enrolled 590 patients who generated 1,298 clinical encounters (590 initial visits and 708 follow-up visits). Monthly visit volume rose during the first program year, peaked at more than 100 visits in mid-2025, and was sustained at roughly 30 to 75 visits per month thereafter (Figure 1). Cumulative enrollment climbed across the 24-month period, reflecting continuous intake of new patients alongside a growing follow-up population.

**Figure 1.**
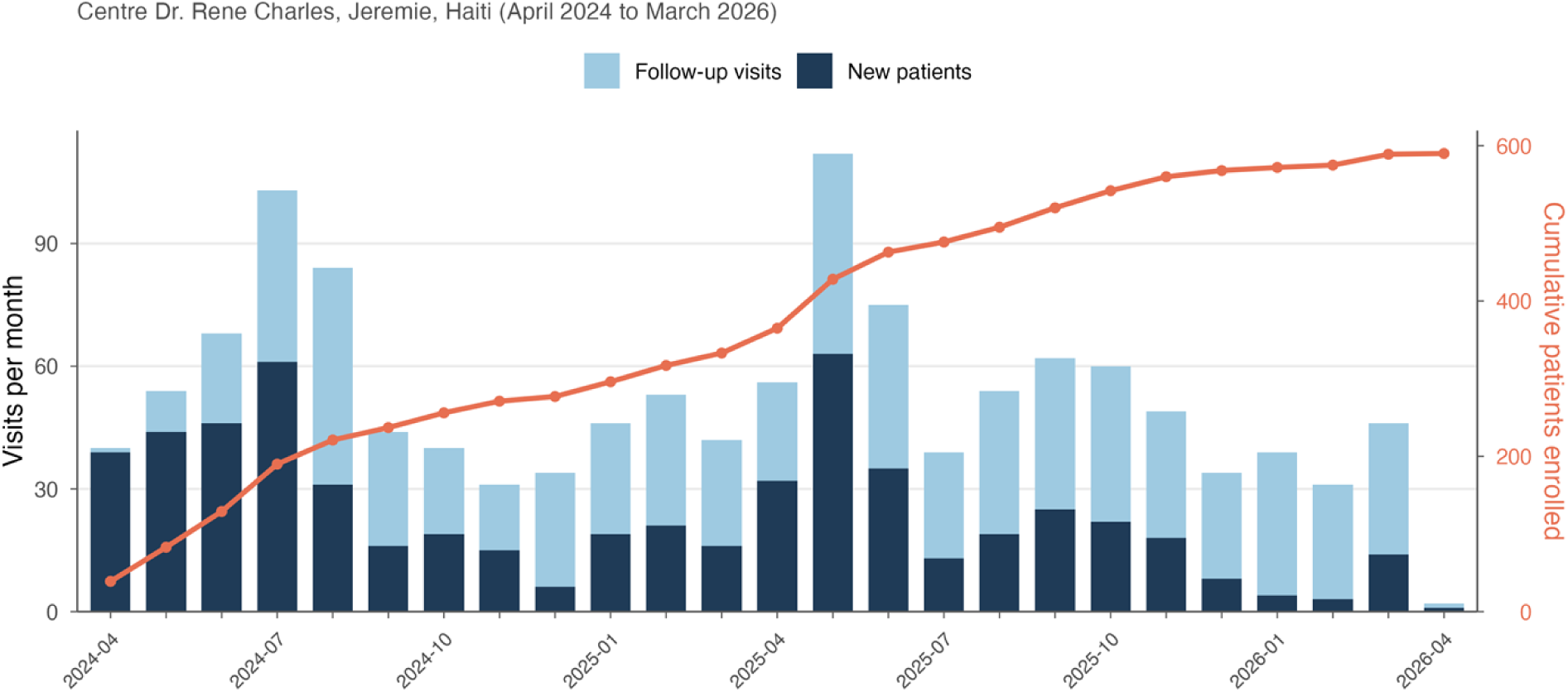
Monthly visit volume (new patients in dark blue, follow-up visits in light blue) and cumulative enrollment (orange line), April 2024 to March 2026.

### Patient Demographics and Clinical Profile

Baseline characteristics are summarized in Table 1. The mean age at enrollment was 58.2 years, and 437 patients (74.1%) were women. Most patients resided in Jérémie (311 of 378, 82.3%), and educational disadvantage was common: 220 of 346 (63.6%) had primary-level education or none. Hypertension alone was the most common diagnosis (312 patients, 52.9%), followed by combined diabetes and hypertension (158 patients, 26.8%) and diabetes without hypertension (120 patients, 20.3%); diabetes of any type was present in 278 patients (47.1%) and hypertension in 470 (79.7%). Among patients with a documented subtype, 125 of 147 (85.0%) had type 2 disease. At the first visit, mean systolic and diastolic pressures were 142.8 and 90.2 mmHg, and only 187 of 583 patients (32.1%) met the control target. Mean BMI was 24.7 kg/m², and abdominal obesity was present in 260 of 402 women (64.7%) and 23 of 144 men (16.0%).

**Table 1.** Baseline Demographic and Clinical Characteristics (N = 590).

| Characteristic | Value | N |
| --- | --- | --- |
| Age, mean ± SD, yr | 58.2 ± 14.6 | 590 |
| Female sex, n (%) | 437 (74.1) | 590 |
| Residence in Jérémie, n (%) | 311 (82.3) | 378 |
| No formal education, n (%) | 102 (29.5) | 346 |
| Primary education only, n (%) | 118 (34.1) | 346 |
| Primary diagnosis, n (%): |  |  |
| Hypertension only (HT) | 312 (52.9) | 590 |
| Diabetes and hypertension (DT) | 158 (26.8) | 590 |
| Diabetes mellitus only (DM) | 120 (20.3) | 590 |
| Type 2 diabetes (among subtyped) | 125 (85.0) | 147 |
| History of hypertension, n (%) | 311 (82.9) | 375 |
| History of diabetes, n (%) | 155 (41.1) | 377 |
| Systolic BP, mean ± SD, mmHg | 142.8 ± 27.0 | 583 |
| Diastolic BP, mean ± SD, mmHg | 90.2 ± 15.7 | 583 |
| BP controlled at first visit, n (%) | 187 (32.1) | 583 |
| BMI, mean ± SD, kg/m<sup>2</sup> | 24.7 ± 5.1 | 583 |
| Waist ≥88 cm (women), n (%) | 260 (64.7) | 402 |
| Waist ≥102 cm (men), n (%) | 23 (16.0) | 144 |
| Current tobacco use, n (%) | 29 (4.9) | 588 |
| Current alcohol use, n (%) | 90 (15.3) | 588 |

### Diagnostic Coverage

Blood pressure and BMI were recorded at virtually every encounter (98.8% and 99.3% of patients; Table 2). Fasting glucose was measured at least once in 315 patients (53.4%) and HbA1c in 190 of 278 patients with diabetes (68.3%). Coverage of investigations requiring reagents or specialized equipment was lower (total cholesterol, 28.0%; urine ACR, 30.7%; serum creatinine or eGFR, 17.1%; electrocardiography, 12.1% of patients with hypertension; and fundoscopy, 3.6% of patients with diabetes), because testing was guided by availability and clinical priority rather than performed uniformly. Prevalence estimates that follow therefore apply to the tested subgroups.

**Table 2.** Diagnostic Coverage, According to Investigation.

| Investigation | Measurements | Patients tested | Eligible | Coverage |
| --- | --- | --- | --- | --- |
| **Blood pressure** | 1,264 | 583 | 590 | 98.8% |
| **Body-mass index** | 1,289 | 586 | 590 | 99.3% |
| **Fasting glucose** | 646 | 315 | 590 | 53.4% |
| **Glycated hemoglobin** | 227 | 190 | 278 (DM/DT) | 68.3% |
| **Total cholesterol** | 181 | 165 | 590 | 28.0% |
| **Serum creatinine / eGFR** | 113 | 101 | 590 | 17.1% |
| **Urine ACR** | 193 | 181 | 590 | 30.7% |
| **Electrocardiography** | 58 | 57 | 470 (HT/DT) | 12.1% |
| **Fundoscopy** | 10 | 10 | 278 (DM/DT) | 3.6% |

eGFR denotes estimated glomerular filtration rate and ACR albumin-to-creatinine ratio. Eligible denominators: all 590 patients for general screening; 278 patients with diabetes (DM or DT) for HbA1c and fundoscopy; and 470 patients with hypertension (HT or DT) for electrocardiography.

### Blood-Pressure and Glycemic Outcomes

In a linear mixed-effects model fit to all patients with hypertension, systolic blood pressure declined over calendar time by 0.78 mmHg per month (95% CI, 0.36 to 1.21; P=0.005) and diastolic pressure by 0.64 mmHg per month (95% CI, 0.36 to 0.92; P<0.001); because a time slope can be estimated only from patients seen more than once, this longitudinal estimate is informed by the 210 hypertensive patients with two or more measurements (470 patients contributed 1,010 measurements overall; Table 3 and Figure 2A). The paired first-to-last analysis, restricted to the 210 patients with hypertension who had at least two blood-pressure measurements, was concordant and somewhat larger: mean systolic pressure fell by 9.0 mmHg, from 149.4 to 140.4 mmHg (95% CI, 5.7 to 12.2; P<0.001), and diastolic pressure fell by 6.9 mmHg (95% CI, 4.8 to 9.0; P<0.001), over a median of 47 days. The proportion of these patients with controlled blood pressure rose from 21.4% to 38.1% (P<0.001 by McNemar’s test), with 48 patients moving from uncontrolled to controlled status and 13 moving in the opposite direction.

**Figure 2.**
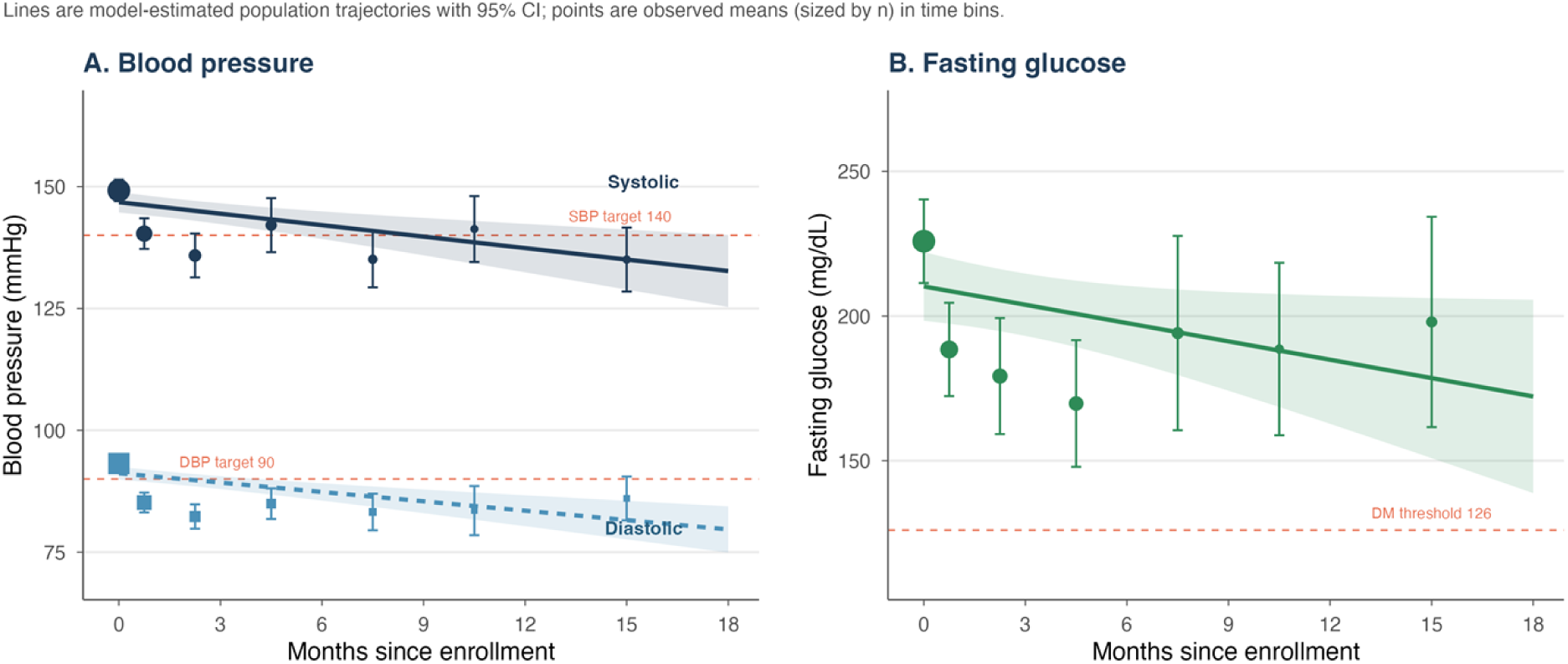
Cardiometabolic trends over calendar time. Solid and dashed lines are population trajectories estimated by linear mixed-effects models with 95% confidence bands; points are observed means (sized by the number of contributing measurements) within time bins. Panel A shows systolic and diastolic blood pressure among patients with hypertension; Panel B shows fasting glucose among patients with diabetes. Dashed horizontal lines mark clinical thresholds.

**Table 3.** Change in Cardiometabolic Outcomes over Calendar Time (Linear Mixed-Effects Models).

| Outcome (population) | Patients | Measurements | Change per month (95% CI) | P value |
| --- | --- | --- | --- | --- |
| Systolic BP (hypertension) | 470 | 1,010 | -0.78 (-1.21 to -0.36) | 0.005 |
| Diastolic BP (hypertension) | 470 | 1,010 | -0.64 (-0.92 to -0.36) | <0.001 |
| Fasting glucose (diabetes) | 224 | 551 | -2.11 (-4.15 to -0.07) | 0.05 |

Estimates are fixed-effect slopes for time (months since enrollment) from linear mixed-effects models with a random intercept and random slope per patient, using all available measurements. BP denotes blood pressure. Change per month is expressed in mmHg for blood pressure and mg/dL for fasting glucose; *P* values use the Satterthwaite approximation.

Among the 224 of 278 patients with diabetes who had a fasting glucose measurement, fasting glucose also declined over time, by 2.1 mg/dL per month in the mixed-effects model (95% CI, 0.1 to 4.2; *P*=0.05; 551 measurements; Figure 2B), a more modest and less certain trend than for blood pressure. The paired analysis of the 115 patients with diabetes who had at least two fasting-glucose measurements showed a larger first-to-last fall of 52.5 mg/dL, from 236.5 to 184.0 mg/dL (95% CI, 33.4 to 71.6; *P*<0.001), over a median of 84 days. Durable glycemic control, however, remained poor: across 190 patients with at least one HbA1c measurement (227 measurements), the mean was 10.4%, with 57.7% of values at or above 10% and only 13.2% below the 7% target.

### End-Organ Damage Among Tested Patients

Targeted screening, performed in only a small and clinically selected fraction of the cohort, frequently detected end-organ damage (Figure 3). These investigations were ordered at the clinician’s discretion, typically when a complication was already suspected, and were completed for a minority of patients (urine ACR in 181 of 590 patients, 30.7%; electrocardiography in 57 of 470 patients with hypertension, 12.1%; serum creatinine in 101 of 590, 17.1%; and fundoscopy in 10 of 278 patients with diabetes, 3.6%). The proportions below are therefore specific to this tested subgroup and greatly overestimate the prevalence that would be found if the whole cohort were screened; they should not be read as the baseline prevalence of end-organ disease in the clinic population. Among those tested, albuminuria (ACR of at least 30 mg/g) was present in 170 of 193 measurements (88.1%; 95% CI, 82.8 to 91.9; 181 patients), abnormal electrocardiograms in 36 of 57 patients with hypertension (63.2%; 95% CI, 50.2 to 74.5), and reduced eGFR (below 60) in 27 of 101 patients (26.7%; 95% CI, 19.1 to 36.1). Fundoscopy was abnormal in 8 of 10 patients examined, which is best read as a signal of unmet retinal screening need rather than a prevalence estimate.

**Figure 3.**
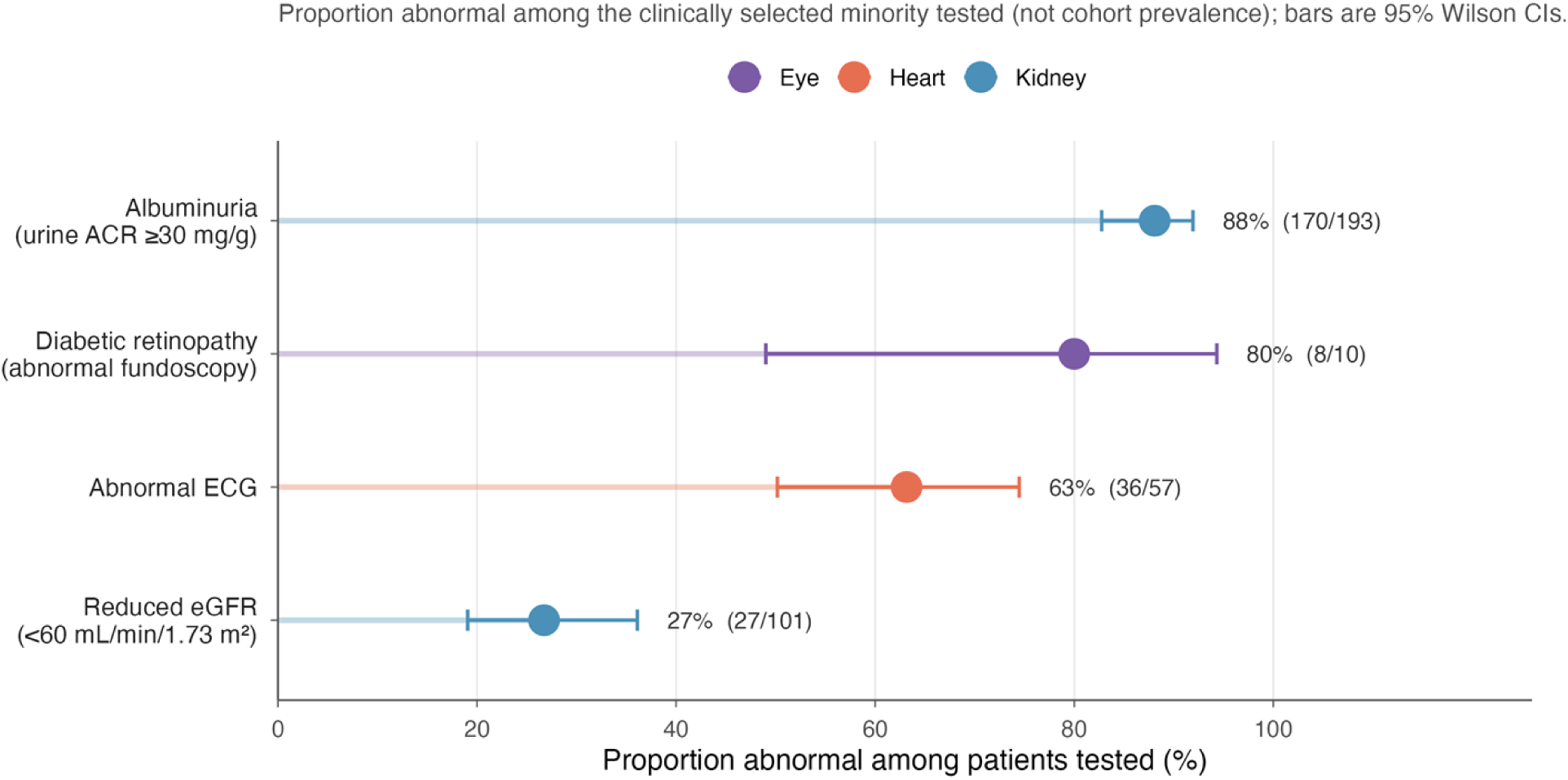
End-organ damage detected among the clinically selected patients who underwent screening. Points indicate the proportion abnormal among tested patients and horizontal bars 95% Wilson confidence intervals; denominators are the numbers tested (a minority of the cohort), so these proportions overestimate cohort-wide prevalence.

### Retention and Follow-up

Patients attended a mean of 2.2 visits (range, 1 to 23). A total of 269 patients (45.6%) returned for at least two visits, 136 (23.1%) for three or more, and 65 (11.0%) for five or more (Figure 4B). Among returners, the median follow-up was 49 days (interquartile range, 17 to 143). Time-horizon retention was 42.8% at 90 days, 43.7% at 180 days, and 44.7% at 365 days, indicating that attrition reflected early disengagement rather than insufficient observation time. Returning and single-visit patients were similar in age, sex, and baseline blood pressure (Table 4); the principal difference was diagnostic, with patients carrying dual diabetes and hypertension more likely to return than those with hypertension alone (about 63% vs. 36%), and hypertension alone predominating among single-visit attenders. At the most recent visit, 26.3% of patients with diabetes were receiving insulin and 65.1% an oral antidiabetic agent.

**Figure 4.**
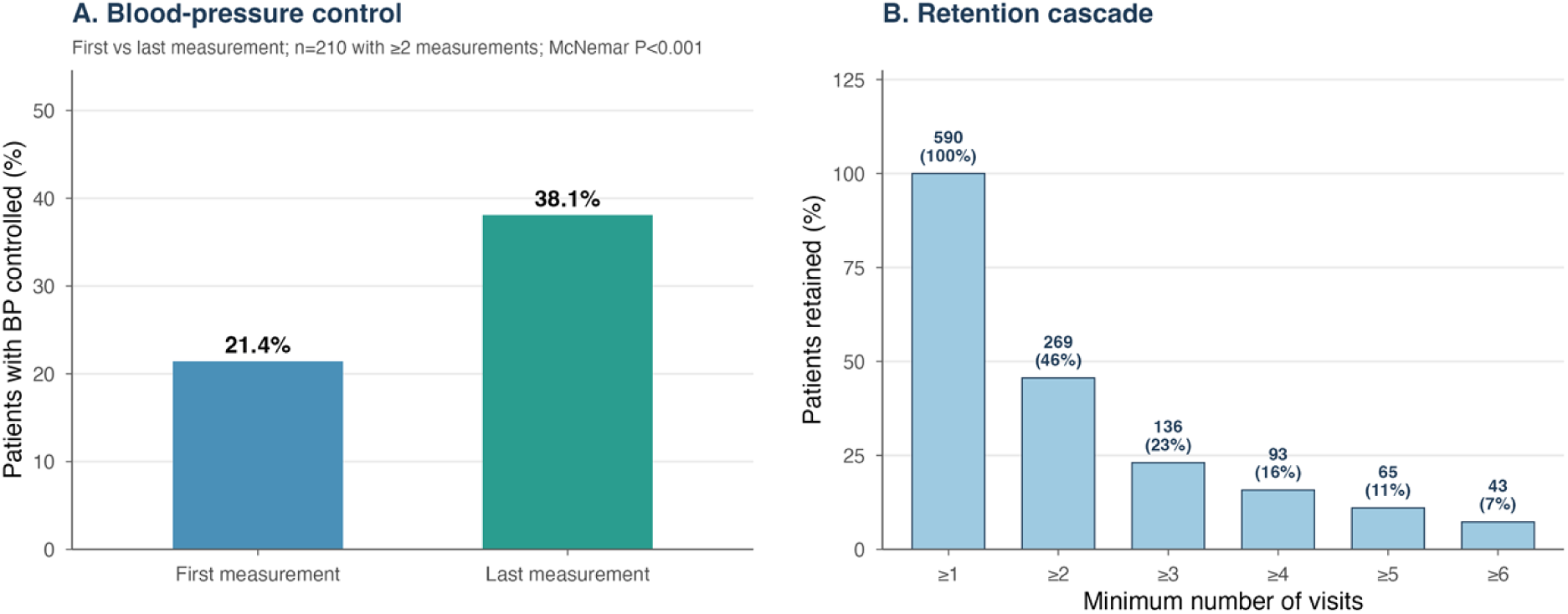
Summary outcomes. Panel A shows blood-pressure control at the first versus last measurement among the 210 patients with hypertension who had at least two measurements (McNemar *P*<0.001); Panel B shows the patient-retention cascade by visit threshold.

**Table 4.**
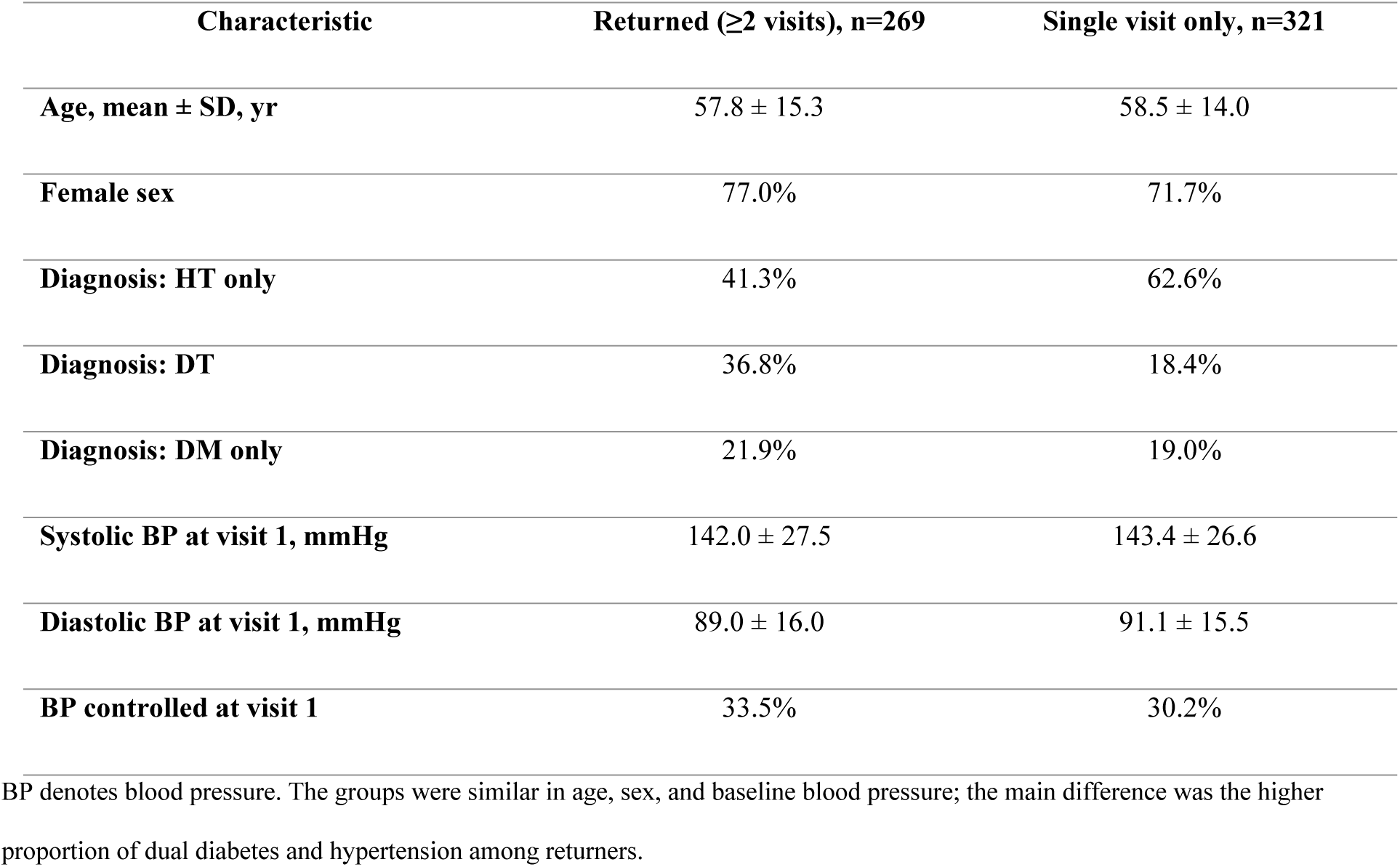
Baseline Characteristics of Returning versus Single-Visit Patients.

| Characteristic | Returned ($\geq 2$ visits), n=269 | Single visit only, n=321 |
| --- | --- | --- |
| Age, mean ± SD, yr | 57.8 ± 15.3 | 58.5 ± 14.0 |
| Female sex | 77.0% | 71.7% |
| Diagnosis: HT only | 41.3% | 62.6% |
| Diagnosis: DT | 36.8% | 18.4% |
| Diagnosis: DM only | 21.9% | 19.0% |
| Systolic BP at visit 1, mmHg | 142.0 ± 27.5 | 143.4 ± 26.6 |
| Diastolic BP at visit 1, mmHg | 89.0 ± 16.0 | 91.1 ± 15.5 |
| BP controlled at visit 1 | 33.5% | 30.2% |

BP denotes blood pressure. The groups were similar in age, sex, and baseline blood pressure; the main difference was the higher proportion of dual diabetes and hypertension among returners.

## DISCUSSION

This study reports the first 24 months of a dedicated chronic disease clinic for diabetes and hypertension in the Grand’Anse Department of Haiti. In a region that previously had no registry, screening protocol, or follow-up system for noncommunicable disease, the Centre Dr. René Charles enrolled 590 patients, delivered 1,298 encounters, and generated the first structured longitudinal cardiometabolic data for this population. Two findings define the cohort. A functioning, protocol-driven NCD service was established and sustained where none had existed. Retention was the central challenge, with 54% of patients attending only once. Among the patients who did engage in follow-up, three clinical observations emerged: blood pressure improved; fasting glucose fell but durable glycemic control, as measured by HbA1c, remained poor; and selective screening of clinically suspected patients detected end-organ damage at a high rate.

Blood-pressure outcomes were the strongest. In a mixed-effects model on calendar time, which uses all available readings and accounts for variable visit timing and within-patient correlation, systolic pressure fell by about 0.78 mmHg per month among patients with hypertension over the observed follow-up period. The paired first-to-last comparison gave a directionally concordant but substantially larger estimate (9.0 mmHg), a gap itself consistent with regression to the mean, and blood-pressure control rose from 21.4% to 38.1%, with nearly four times as many patients gaining control as losing it. Because these estimates are drawn only from patients with repeat measurements and are anchored to a single baseline reading, part of the apparent decline may reflect regression to the mean; the mixed-effects estimate is less sensitive to a single high baseline value and is reassuring on this point, but we present the time trend as exploratory rather than as a demonstrated treatment effect. If sustained, a reduction of this magnitude would be clinically meaningful, since each 10 mmHg fall in systolic pressure is associated with roughly a 20% lower risk of major cardiovascular events,^33^ and would be consistent with the dedicated, nurse-led, protocol-based NCD services piloted elsewhere in sub-Saharan Africa, where general primary-care facilities are often unprepared for chronic disease.^34,35^

The glycemic findings were weaker and, in the durable measure, essentially null. Fasting glucose fell substantially in the paired analysis (by 52.5 mg/dL), but this comparison begins from a very high baseline (mean, 236.5 mg/dL), relies on self-reported fasting and point-of-care measurement, and is therefore especially vulnerable to regression to the mean; the calendar-time mixed model showed only a modest decline of about 2 mg/dL per month that merely reached the conventional significance threshold (P=0.05). Mean HbA1c, the durable measure of control, remained 10.4%, with more than half of all values at or above 10% and only 13.2% at target, so we read the glycemic results as showing little durable improvement to date. This gap between a large first-to-last fall, a borderline time trend, and persistently high HbA1c is the central clinical lesson of the cohort. It reflects the severity of hyperglycemia at presentation after years of unmanaged disease, the short follow-up available to date, and a constrained formulary in which insulin supply is intermittent, the range of oral agents is narrow, and newer therapies such as SGLT2 inhibitors and GLP-1 receptor agonists are unavailable. At the most recent visit, 65% of patients with diabetes were taking an oral agent and 26% insulin. Closing the gap between short-term fasting glucose and durable HbA1c control will require a more reliable medication supply and intensified titration, not simply more visits.

Albuminuria was present in 88% and abnormal electrocardiograms in 63% of patients tested, but these investigations were performed in only a small, clinically selected fraction of the cohort (31% for albuminuria, 12% for electrocardiography, and fewer still for fundoscopy), typically when a complication was already suspected. These proportions are therefore best understood as the detection rate of selective, indication-driven testing, and they overstate what the prevalence would be if the whole cohort were screened; they are not estimates of baseline end-organ-disease prevalence in the clinic population. For context, the population-based Haiti Cardiovascular Disease Cohort reported an age-standardized chronic kidney disease prevalence of 14% in Port-au-Prince,^23^ far below the rate in our tested subgroup, exactly as expected when testing is concentrated among higher-risk patients; a high chronic kidney disease burden is likewise reported in general populations across the African continent.^36^ With that caveat, the frequency of abnormal findings across several independent organ systems indicates that treatable, otherwise silent complications are common among the patients tested, and it argues for expanding systematic, protocolized screening so that prevalence can be estimated without selection and more patients with early complications can be identified.

Retention is the clinic’s defining operational challenge. Just under half of patients returned for a second visit, and the time-horizon analysis showed that this reflected early disengagement rather than insufficient follow-up time, with retention near 43 to 45% whether measured at 90, 180, or 365 days. The barriers are those common to NCD programs across low-income settings:^10^ most patients had primary education or less, and travel from remote communities is difficult and costly. Patients with dual diabetes and hypertension were nearly twice as likely to return as those with hypertension alone (about 63% vs. 36%), which suggests that perceived disease severity drives engagement and that patients with asymptomatic single conditions are the group most likely to be lost. This points to clear, testable interventions, including community health-worker follow-up, decentralized medication dispensing, appointment reminders, and transport support, targeted especially at patients with asymptomatic hypertension.

Women made up 74% of the cohort. Although Haitian surveys report somewhat higher NCD prevalence in women,^19^ an imbalance of this degree more likely reflects health-seeking behavior; abdominal obesity was present in 65% of women versus 16% of men, a marked concentration of metabolic risk in the female patients. The under-representation of men is itself a concern, because men in such settings often present later and use health services less, and workplace and community strategies to engage them should be built into future program design.

This study has several limitations. As a single-center cohort without a control group, the observed improvements are associations rather than proven treatment effects, and we report no hard endpoints such as mortality or cardiovascular events. Because baseline blood pressure was recorded from a single seated reading, part of the apparent first-to-last decline may reflect regression to the mean; the calendar-time mixed-effects model, which is less sensitive to a single extreme baseline value and uses all observations, mitigates but does not eliminate this concern. Follow-up was short (median 49 days among returners) and attrition was high, so the longitudinal estimates rest disproportionately on the engaged minority and may not generalize to all enrolled patients. Most patients resided in or near Jérémie town, so the cohort may under-represent the more remote communities across the department that the clinic ultimately aims to serve. Diagnostic testing was incomplete and strongly selective, so the complication proportions apply to a small tested subgroup and overestimate cohort-wide prevalence; the fundoscopy figure, from only 10 patients, is illustrative rather than quantitative. Fasting status for glucose was self-reported, and medication-adherence questionnaires were collected but not analyzed.^37^ These constraints temper our conclusions but do not negate the core observations: a functioning chronic disease service was established, engaged patients improved, and selective screening uncovered a high burden of previously undetected complications.

These data establish that a dedicated chronic disease facility can be built in one of the world’s most resource-limited environments, drawing on the delivery model developed for HIV care in the same region.^38^ The remaining requirement is sustained political and financial commitment.

## CONCLUSIONS

The Centre Dr. René Charles in Jérémie is the first dedicated chronic disease facility in the Grand’Anse Department. In 24 months it enrolled 590 patients and delivered 1,298 encounters, demonstrating that structured NCD care is feasible in a fragile, severely under-resourced setting. Retention, however, was the defining challenge: 54% of patients attended only once. Among the subset who returned, blood pressure improved in both mixed-effects and paired analyses (systolic 0.78 mmHg per month, P=0.005; 9.0 mmHg first-to-last, P<0.001; control rising from 21.4% to 38.1%, P<0.001), a promising but exploratory signal that will require controlled, longer follow-up to confirm. Glycemic control showed little durable change: fasting glucose fell but the calendar-time trend was borderline, and persistently elevated HbA1c (mean, 10.4%) marks the limit of what the current formulary and follow-up interval can achieve and defines the next clinical priority.

Indication-driven screening of clinically suspected patients frequently detected end-organ damage, pointing to a likely burden of treatable complications, although the small tested minority overstates cohort-wide prevalence. The priorities now are concrete: expand the pharmacological formulary, broaden and systematize laboratory and screening coverage so prevalence can be estimated without selection, and address the 54% single-visit attrition through community health-worker follow-up, decentralized dispensing, and transport support. At the system level, the Grand’Anse Department needs a reliable NCD medication supply chain, nurse-led care protocols, and routine NCD screening at every primary care contact.^39,40^ These 590 patients are the first cohort to receive structured chronic disease care in the department; the model is feasible, the clinical need is severe, and the case for sustained investment is now supported by data.

## DATA AVAILABILITY

The minimal, de-identified dataset underlying all figures and summary statistics reported in this article (aggregate values only, containing no individual records) is publicly available in the Zenodo repository at https://doi.org/10.5281/zenodo.20915947 (CC BY 4.0). The complete analysis and figure code, written in R, is provided as Supporting Information (S2 File and S3 File); the public dataset reproduces all reported summary statistics and figure values, while reproduction of the individual-level models additionally requires the patient-level clinical registry. That registry cannot be shared publicly because it contains potentially identifying information on a small rural cohort; de-identified patient-level data are available to qualified researchers who meet the criteria for access to confidential clinical data, on request to the FHADIMAC Research Ethics Board, a non-author institutional data-access body.

## ACKNOWLEDGMENTS

The authors thank the clinical staff of the Centre Dr. René Charles for their dedication to patient care, and the community health workers who supported recruitment and follow-up. Special thanks to Joseph Ange Dany, Edel Marie Geralde, Antoine Michel Mie Elisabeth, Jeanty Josue, Philippe Kendy, Samuel Beatrice, Franck Candys Nahomie, Juste Donal, Jean Celome. We are grateful to the patients who entrusted the clinic with their care.

## SUPPORTING INFORMATION

**S1 File.** Pre-specified statistical analysis plan.

**S2 File.** Analysis code (R).

**S3 File.** Figure code (R).

